# Synergistic associations of cognitive and motor impairments with functional outcome in covert cerebral small vessel disease

**DOI:** 10.1101/2021.05.20.21257306

**Authors:** Hanna Jokinen, Hanna M. Laakso, Matti Ahlström, Anne Arola, Juha Lempiäinen, Johanna Pitkänen, Teemu Paajanen, Sietske A. M. Sikkes, Juha Koikkalainen, Jyrki Lötjönen, Antti Korvenoja, Timo Erkinjuntti, Susanna Melkas

## Abstract

**Objective:** Cognitive and motor impairments are the key clinical manifestations of cerebral small vessel disease (SVD), but their interrelations and combined effects on functional outcome have not been elucidated. We investigated the associations between cognitive and motor functions and their interactions and mediating effects on instrumental activities of daily living (IADL) and quality of life in older individuals with various degrees of white matter hyperintensities (WMH).

**Methods:** Participants of the Helsinki Small Vessel Disease Study (n=152) were assessed according to an extensive clinical, neuropsychological and MRI protocol. Cognitive composite scores for global cognition, processing speed, executive functions and memory were constructed from multiple tests within each domain. Physical examination included measures of gait speed, balance (single-leg-stance) and functional mobility (timed-up-and-go test). IADL was evaluated with a proxy-based Amsterdam IADL questionnaire and quality of life with a self-report EUROHIS-Qol index. Volumes of WMH and gray matter (GM) were obtained with automated segmentation. Sets of linear regression analyses were used to model the associations between motor and cognitive performances, WMH and GM volumes, and IADL and quality of life.

**Results:** Domain-specific cognitive and motor functions had strong interrelations with each other, and they were significantly associated with IADL, quality of life as well as WMH and GM volumes. A consistent pattern on significant interactions between cognitive and motor functions was found on IADL, but not on quality of life. In particular, low cognitive scores together with decline in the timed-up-and-go test and gait speed were strongly related to impaired IADL. The association of WMH volume with IADL was mediated by global cognition, whereas the association of GM volume with IADL was mediated by global cognition and timed up-and-go performance.

**Conclusion:** The results highlight the complex interplay and synergism between motor and cognitive abilities on functional outcome in SVD. The combined effect of motor and cognitive disturbances on IADL is likely to be greater than the individual effects of each of the two impairments. WMH and brain atrophy contribute to disability through cognitive and motor impairment.

## INTRODUCTION

Cognitive and motor impairments become increasingly prevalent with aging and pose a major public health concern in older people. Cognitive abilities play a key role in regulating complex motor functions such as gait and balance. Cognitive impairment, executive dysfunction in particular, has been associated with risk of falls in community-dwelling older individuals.^1^ Cerebral small vessel disease (SVD) is the leading cause of vascular cognitive impairment and a major contributor to functional disability.^2, 3^ It is characterized by insidious onset, gradually accumulating brain changes and progressive clinical symptoms.^3, 4^ Among the characteristic neuroimaging findings of SVD, white matter hyperintensities (WMH), lacunar infarcts and brain atrophy have been the strongest determinants of cognitive decline.^5-7^ Executive functions and processing speed have been regarded as the core domains of cognitive impairment in the early stages of SVD, but eventually the deterioration becomes more global and affects a wide range of cognitive functions.^8, 9^

Along with cognitive impairment, decline in motor functions is another important adverse outcome of SVD. The extent of WMH has been associated with impaired gait and balance as well as history of falls,^10-14^ whereas measures of gray matter (GM) atrophy have been related to gait performance.^14, 15^ Previously, the cognitive and motor symptoms of SVD have been explored in isolation, and therefore, little is known about their interrelations and combined associations with functional outcome. A recent retrospective study has reported executive functions, but not global cognition, to be associated with risk of falls in patients with chronic cerebrovascular disease, but it did not demonstrate any relationship between measures of cognition and balance.^16^ Another study has found an association between mobility and cognitive performance independently of cerebral lesion burden among older patients with atrial fibrillation.^17^

This study investigated the relationships and functional significance of cognitive and motor abilities in older individuals with different degrees of WMH by using standard physical and neuropsychological assessments and quantitative evaluation of brain changes on structural MRI. Specifically, our objectives were to

a. confirm the associations of domain-specific cognitive functions (global cognition, executive functions, processing speed and memory) and motor performances (gait speed, balance and functional mobility) with WMH and GM volumes
b. investigate the reciprocal relationships between different motor and cognitive functions
c. show the associations and interactions of cognitive and motor functions on instrumental activities in daily living (IADL) and quality of life
d. explore the extent to which cognitive and motor functions mediate the effect of WMH and GM volumes on functional outcome.

## METHODS

### Participants

In total, 152 subjects with different degrees of WMH were enrolled in the Helsinki Small Vessel Disease Study, a prospective cohort study designed to investigate the imaging, clinical and cognitive characteristics of cerebral SVD. Subjects who had recently undergone brain scanning for one reason or another (table 1) were recruited from the imaging registry of the Helsinki University Hospital, Finland between October 2016 and March 2020.

**Table 1.**
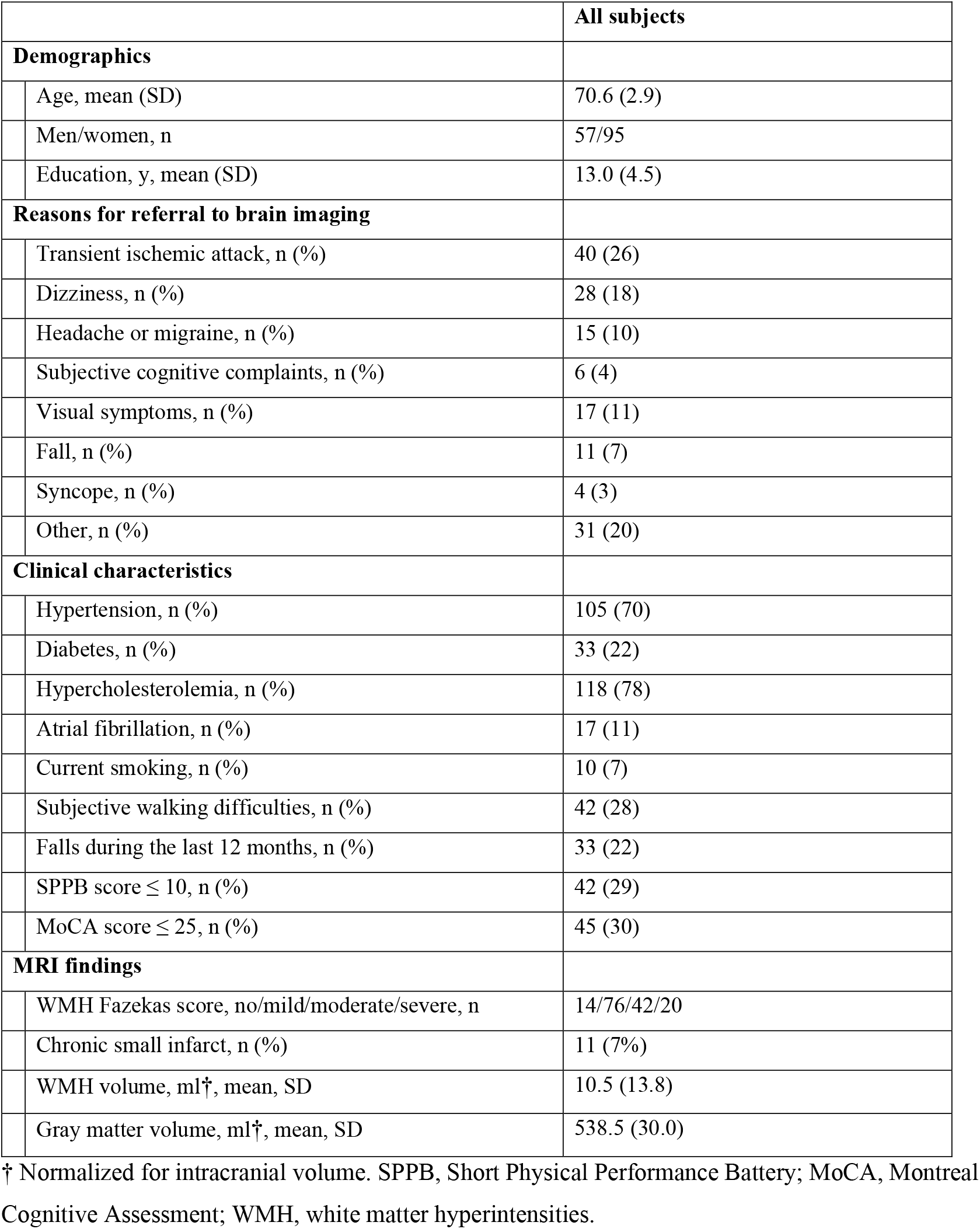
Characteristics of the subjects in the Helsinki Small Vessel Disease Study (n=152)

|  |  | All subjects |
| --- | --- | --- |
| **Demographics** |  |  |
|  | Age, mean (SD) | 70.6 (2.9) |
|  | Men/women, n | 57/95 |
|  | Education, y, mean (SD) | 13.0 (4.5) |
| **Reasons for referral to brain imaging** |  |  |
|  | Transient ischemic attack, n (%) | 40 (26) |
|  | Dizziness, n (%) | 28 (18) |
|  | Headache or migraine, n (%) | 15 (10) |
|  | Subjective cognitive complaints, n (%) | 6 (4) |
|  | Visual symptoms, n (%) | 17 (11) |
|  | Fall, n (%) | 11 (7) |
|  | Syncope, n (%) | 4 (3) |
|  | Other, n (%) | 31 (20) |
| **Clinical characteristics** |  |  |
|  | Hypertension, n (%) | 105 (70) |
|  | Diabetes, n (%) | 33 (22) |
|  | Hypercholesterolemia, n (%) | 118 (78) |
|  | Atrial fibrillation, n (%) | 17 (11) |
|  | Current smoking, n (%) | 10 (7) |
|  | Subjective walking difficulties, n (%) | 42 (28) |
|  | Falls during the last 12 months, n (%) | 33 (22) |
| | SPPB score $\leq 10$, n (%) | 42 (29) |
| | MoCA score $\leq 25$, n (%) | 45 (30) |
| **MRI findings** |  |  |
|  | WMH Fazekas score, no/mild/moderate/severe, n | 14/76/42/20 |
|  | Chronic small infarct, n (%) | 11 (7%) |
|  | WMH volume, ml<sup>†</sup>, mean, SD | 10.5 (13.8) |
|  | Gray matter volume, ml<sup>†</sup>, mean, SD | 538.5 (30.0) |

<sup>†</sup> Normalized for intracranial volume. SPPB, Short Physical Performance Battery; MoCA, Montreal Cognitive Assessment; WMH, white matter hyperintensities.

At study entry, all subjects underwent clinical, physical and neuropsychological examinations and a subsequent brain MRI carried out at three visits within approximately one month. Detailed data of demographic factors, social background, medical history, cardiovascular risk factors and lifestyle was collected with a structured interview and review of all available medical records.

The inclusion criteria were: a) age 65-75 years at the time of enrollment; b) place of residence within the Helsinki and Uusimaa Hospital District; c) occurrence of not more than minor, temporary and local neurological symptoms (having manifested 3 to 12 months before the enrolment), or no neurological symptoms at all; d) no more than slight disability in basic daily activities as defined by the modified Ranking Scale score 0-2 (able to look after one’s own affairs without assistance); and e)sufficient vision and hearing, and Finnish language skills to complete the full assessment protocol.

The exclusion criteria were: a) significant neurological disease (e.g. symptomatic stroke, multiple sclerosis or treatment resistant epilepsy); b) severe diagnosed psychiatric disorder; c) current substance abuse; d) severe other medical condition preventing participation; e) traumatic brain injury that has required hospitalization; f) severe intellectual disability; and g) inability or refusal to undergo brain MRI. Additional exclusion criteria based on MRI findings were: a) cortical infarct; b) subcortical infarct larger than 15 mm (20 mm on diffusion-weighted images); c) hemorrhage larger than microbleed (over 10 mm); d) brain tumor; and e) contusion, traumatic subarachnoidal or intracranial hemorrhage, distinct diffuse axonal injury, as evaluated by an experienced neuroradiologist.

### Standard Protocol Approvals, Registrations, and Patient Consents

The study was approved by the Ethics Committee of the Helsinki University Hospital and conducted according to the Declaration of Helsinki. Informed written consent was received from each subject.

### Neuroimaging

The subjects were scanned using a 3 T MRI scanner with 32-channel head coil (Siemens Magnetom Skyra/Verio). The protocol included the following sequences: a fast T1 gradient echo localizer in three orthogonal directions (0.9×0.8 mm in-plane resolution, 8 mm slice thickness, TR 9 ms, TE 4 ms, FA 20 degrees), sagittal 3D FLAIR SPACE (1.1×1.0 mm in-plane resolution, 1 mm slice thickness, 176 slices, TR 6000 ms, TE 389, ms, TI 2100 ms), sagittal 3D T2 SPACE (1.0×1.0 mm in-plane resolution, 1 mm slice thickness, 176 slices TR 3200 ms, TE 416 ms), and sagittal 3D T1 MPRAGE (1.1×1.0 mm in-plane resolution, 1 mm slice thickness, 176 slices, TR 1900 ms, TE 2.47 ms, FA 9 degrees).

WMH of presumed vascular origin were defined as hyperintense areas in the white matter without cavitation on FLAIR sequences according to the neuroimaging guidelines of the STandards for ReportIng Vascular changes on nEuroimaging (STRIVE).^4^ WMH were first evaluated visually with the modified 4-point Fazekas scale (none, mild, moderate, severe) by an experienced neuroradiologist.^18^ Chronic small infarctions (other than defined in the exclusion criteria) were evaluated by number and location.

Volumes of brain structures and WMH were measured using the fully-automated cNeuro image quantification tool (www.cneuro.com/cmri/ Combinostics Ltd, Finland).^19^ The tool segments the T1 images into 133 brain regions with a multi-atlas method based on 79 manually segmented atlases.^20^ The segmentation of WMH was carried out on FLAIR images according to a multi-stage method based on the Expectation-Maximization algorithm.^21^ All volumes were normalized for intracranial volume.^22^ For the present analyses, the total volumes of WMH and cerebral GM were included. WMH volume was log transformed due to positively skewed distribution.

### Evaluation of motor functions

The evaluation of motor functions included the Short Physical Performance Battery (SPPB), timed up-and-go (TUG) test as well as additional measurements of gait speed and balance. The SPPB is an assessment tool of lower extremity function and mobility comprising of subtests for standing (side-by-side, semitandem, tandem), 4-meter walking and repeated chair stands.^23^ The SPPB composite score ranges from 0 to 12, and score ≤10 was considered impaired in the present analyses. The TUG test is a measure of functional mobility, in which the subject is asked to stand up from an armchair, walk 3 meters, turn, walk back to the chair and sit down again.^24^ The variable used was the time taken in seconds to complete the test. Gait speed was measured from subjects walking an 8-meter course at one’s usual pace. The faster of two trials was recorded (meters/second).^10^ Single-leg stance time was measured by asking the subject to balance on one leg as long as possible with hands on the hips. The best time of four trials (two trials on each leg) with a maximum of 60 seconds was used for the analyses.^10^ In addition, subjective walking difficulties as reported by the patient or informant (yes vs. no) and the occurrence of falls within the last 12 months were recorded.

### Neuropsychological assessment

Neuropsychological examination was administered by using an extensive battery of standard tests to evaluate all key domains of cognitive functions. The tests and variables with references are listed in the Supplementary Table 1. Briefly, processing speed was evaluated with Wechsler Adult Intelligence Scale IV (WAIS-IV) Coding, Stroop test colour-congruent part and Flexible Attention Test (FAT) Numbers (a computerized modification of the Trail Making test A). Executive functions were assessed with the Stroop test colour-incongruent part, Hayling Sentence Completion Test, Brixton Spatial Anticipation Test, verbal fluency and FAT Number-Letter (a computerized modification of the Trail Making B). Working memory was evaluated with Wechsler Memory Scale III (WMS-III) Letter-Number Sequencing and Digit Span, and FAT Visuospatial Span. Memory and learning was assessed with WMS-III Word Lists and Logical memory, and Rey Complex Figure Test (recall). Visuospatial perception was evaluated with WAIS-IV Block Design and Rey Complex Figure Test (copy). Verbal reasoning was assessed with WAIS-IV Similarities.

The distribution of each raw score was checked for outliers and non-normality and a square root or log transformation was applied where appropriate. There were a few missing values, which were not replaced. To construct clinically relevant domain scores, the raw scores were first converted into z scores and scales inverted, if necessary, so that higher values represented better performance in all variables. The domain scores were then calculated by averaging the z scores within each domain taking into account all available data. A global cognition score was defined as the mean of all 6 standardized domain scores. For the present analysis, we included global cognition, executive functions, processing speed and memory scores as the main cognitive composite measures. In addition, Montreal Cognitive Assessment (MoCA), was used as a measure of general cognitive status for descriptive purposes (table 1).^25^

### Evaluation of quality of life and functional abilities

Quality of life was assessed with self-evaluation using the EUROHIS-Qol index, a shortened version of the World Health Organization Quality of Life Instrument WHOQOL-BREF, consisting of eight questions (satisfaction to overall quality of life, health, energy, finances, daily activities, esteem, relationships and living conditions).^26^ The responses are given on a 5-point Likert scale and the total score is calculated by dividing the sum of all responses by the number of valid responses (≥7 required).

Abilities in everyday activities were evaluated by the subject’s informant (spouse, close relative or friend) with the short version of the Amsterdam Instrumental Activities of Daily Living (A-IADL) questionnaire.^27^ The A-IADL questionnaire has underwent comprehensive construct validation and has demonstrated to provide valuable measurement across different countries.^28^ The short A-IADL questionnaire includes 30 items assessing difficulty with a wide range of activities (household, appliances, handling finances, work, computer, leisure activities). Items are scored on a range from 0 (no difficulty) to 4 (unable to perform the activity), in which current performance is compared to past performance. Total scores are calculated based on item response theory and higher scores indicate better functioning.

### Statistical analyses

Data were analyzed using SPSS version 25. Cognitive scores, gait speed, TUG, single-leg stance, EUROHIS-Qol, A-IADL questionnaire, WMH volume and GM volume were analyzed as continuous variables, whereas SPPB, recent falls and subjective walking difficulties were analyzed as dichotomous variables. Complete data for each subject was available for all cognitive composite scores, MRI measures, falls and subjective walking difficulties, while there were 2 to 4 sporadic missing observations in SPPB, TUG, single-leg stance, gait speed and EUROHIS-Qol. Data of the proxy-based A-IADL questionnaire was available for 132 patients, who as a group did not differ from those with missing data (n=20) in age, education, or WMH and GM volumes (p>0.5), but they were more often men (Pearson χ^2^ =5.0, p=0.026). No imputation of missing data was done.

The relationships between MRI measures, motor performances and cognitive scores were analyzed with separate logistic regression models for the dichotomous dependent variables and with linear regression models for the continuous variables. Further analyses focused on the continuous variables only. Interaction terms were added to the linear regression models to examine the combined effects of motor and cognitive functions on quality of life and IADL. Finally, mediation analyses were carried out with the PROCESS 3.5 for SPSS to examine the extent to which cognitive and motor functions explain the association between WMH and functional outcome.^29^ All models were adjusted for age, sex and years of education. Statistical significance was set at 0.05 and false discovery rate (FDR) method was used as the multiplicity correction within each set of analyses (tables 2-5).

**Table 2.** Associations of the total volumes of cerebral white matter hyperintensities (WMH) and gray matter (GM) with motor and cognitive functions

|  | WMH volume | GM volume |
| --- | --- | --- |
| **Motor functions** |  |  |
| Gait speed | -0.25 (**0.002**) † | 0.21 (**0.011**) † |
| Single-leg stance | -0.32 (<**0.001**) † | 0.15 (0.059) |
| Timed up-and-go | 0.29 (<**0.001**) † | -0.32 (<**0.001**) † |
| **Cognitive functions** |  |  |
| Global cognition | -0.26 (<**0.001**) † | 0.25 (<**0.001**) † |
| Executive functions | -0.27 (<**0.001**) † | 0.21 (**0.005**) † |
| Processing speed | -0.26 (<**0.001**) † | 0.19 (**0.019**) † |
| Memory | -0.18 (**0.015**) † | 0.25 (<**0.001**) † |

Linear regression analyses (standardized $\beta$ coefficients and p values) with WMH and GM volume as independent variables and cognitive/motor functions as dependent variables. The associations were examined in separate models adjusted for age, sex and years of education. † = significant after false discovery rate correction.

### Data availability

Anonymized data will be shared on request from a qualified investigator.

## RESULTS

### Characteristics of the subjects

Demographic characteristics, referral reasons to initial brain imaging, clinical characteristics and MRI findings of the subjects are described in table 1. Subject’s age (65–75 years) correlated significantly with GM volume (Pearson *r*=0.19, p=0.018), but not with WMH volume (*r*=0.16, p=0.057).

### Associations between MRI volumetrics, motor functions and cognitive performance

After adjusting for age, sex and education, WMH volume was associated with subjective walking difficulties (OR 2.5, CI 95% 1.0–6.3, p=0.047) and impaired SPPB score ≤ 10 (OR 3.0, CI 95% 1.2–7.8, p=0.024), but not with recent falls. GM volume was inversely associated with impaired SPPB (OR 0.99, CI 95% 0.972–0.998, p=0.029), but not with walking difficulties or recent falls. The associations between WMH and GM volumes and the continuous motor and cognitive measures were all significant, except for the relationship between GM and single-leg stance (table 2).

As shown in table 3, cognitive functions were significantly associated with most of the motor performances. The strongest relationships were observed between global cognition and TUG test, and between executive functions and single-leg stance time (Cohen’s f^2^ 0.14-0.17) corresponding roughly to medium effect sizes after adjusting for confounders.^30^

**Table 3.** Relationships between motor and cognitive performances.

|  | **Gait speed** | **Single-leg stance** | **Timed up-and-go** |
| --- | --- | --- | --- |
| Global cognition | 0.38 (<**0.001**) † | 0.35 (<**0.001**) † | -0.40 (<**0.001**) † |
| Executive functions | 0.38 (<**0.001**) † | 0.43 (<**0.001**) † | -0.34 (<**0.001**) † |
| Processing speed | 0.33 (<**0.001**) † | 0.26 (**0.001**) † | -0.37 (<**0.001**) † |
| Memory | 0.26 (**0.003**) † | 0.14 (0.113) | -0.31 (<**0.001**) † |

Linear regression analyses (standardized $\beta$ coefficients and p values) with cognitive functions as independent variables and motor functions as dependent variables in separate models. All analyses were adjusted for age, sex and years of education. † = significant after false discovery rate correction.

### MRI, cognitive and motor predictors of quality of life and disability

Both WMH and GM volumes were significantly related to informant-evaluated IADL, but not with self-evaluated quality of life (table 4). Nearly all motor and cognitive measures were associated with quality of life and IADL (table 4). Quality of life was most strongly predicted by the TUG test (Cohen’s f^2^ 0.19), whereas IADL was most strongly predicted by global cognition and the TUG test (Cohen’s f^2^ 0.12 and 0.11, respectively).

**Table 4.** Associations of motor and cognitive functions with quality of life and instrumental activities of daily living (IADL)

|  | Quality of life | IADL |
| --- | --- | --- |
| **MRI measures** |  |  |
| WMH volume | -0.16 (0.053) | -0.28 (**0.001**) † |
| GM volume | 0.10 (0.229) | 0.24 (**0.009**) † |
| **Motor functions** |  |  |
| Gait speed | 0.29 (<**0.001**) † | 0.26 (**0.004**) † |
| Single-leg stance | 0.19 (**0.041**) † | 0.23 (**0.016**) † |
| Timed up-and-go | -0.43 (<**0.001**) † | -0.35 (<**0.001**) † |
| **Cognitive functions** |  |  |
| Global cognition | 0.25 (**0.007**) † | 0.37 (<**0.001**) † |
| Executive functions | 0.24 (**0.008**) † | 0.24 (**0.011**) † |
| Processing speed | 0.16 (0.067) | 0.31 (<**0.001**) † |
| Memory | 0.19 (**0.035**) † | 0.27 (**0.006**) † |

Linear regression analyses (standardized $\beta$ coefficients and p values) with MRI measures and cognitive/motor functions as independent variables and quality of life (self-evaluation) and IADL (informant-evaluation) as dependent variables. The associations were examined in separate models adjusted for age, sex and years of education. † = significant after false discovery rate correction.

We further examined the combined effects of motor and cognitive functions on quality of life and IADL by adding the centered predictor variables and their interaction terms in the models adjusted for the demographic factors. We found significant interactions between several cognitive and motor functions on IADL suggesting synergistic effects between these two factors on functional outcome, but only one (memory × gait speed) on quality of life, which did not survive the FDR correction (table 5). In particular, low cognitive scores together with decline in the TUG test and gait speed were strongly related to impaired IADL. As can be seen in figure 1, subjects with low performance in both cognitive and motor assessments had disproportionally low scores in IADL, while either impairment alone had little impact.

**Table 5.** Interactions between cognitive and motor functions on quality of life and instrumental activities of daily living (IADL)

|  | Quality of life | IADL |
| --- | --- | --- |
| Global cognition × gait speed | -0.12 (0.159) | -0.27 (**0.001**) † |
| Global cognition × single-leg stance | -0.10 (0.264) | -0.11 (0.200) |
| Global cognition × timed up-and-go | 0.15 (0.068) | 0.27 (**0.001**) † |
| Executive functions × gait speed | -0.09 (0.286) | -0.25 (**0.004**) † |
| Executive functions × single-leg stance | -0.08 (0.363) | -0.14 (0.108) |
| Executive functions × timed up-and-go | 0.09 (0.274) | 0.19 (**0.028**) † |
| Processing speed × gait speed | -0.09 (0.311) | -0.29 (**<0.001**) † |
| Processing speed × single-leg stance | -0.11 (0.184) | -0.19 (**0.031**) † |
| Processing speed × timed up-and-go | 0.14 (0.110) | 0.34 (**<0.001**) † |
| Memory × gait speed | -0.19 (**0.019**) | -0.31 (**<0.001**) † |
| Memory × single-leg stance | -0.10 (0.221) | -0.19 (**0.028**) † |
| Memory × timed up-and-go | 0.14 (0.088) | 0.33 (**<0.001**) † |

Linear regression results (standardized $\beta$ coefficients and p values) for the interaction terms of cognitive and motor functions on quality of life and IADL. The interactions were examined adjusted for age, sex and years of education. † = significant after false discovery rate correction.

**Figure 1.**
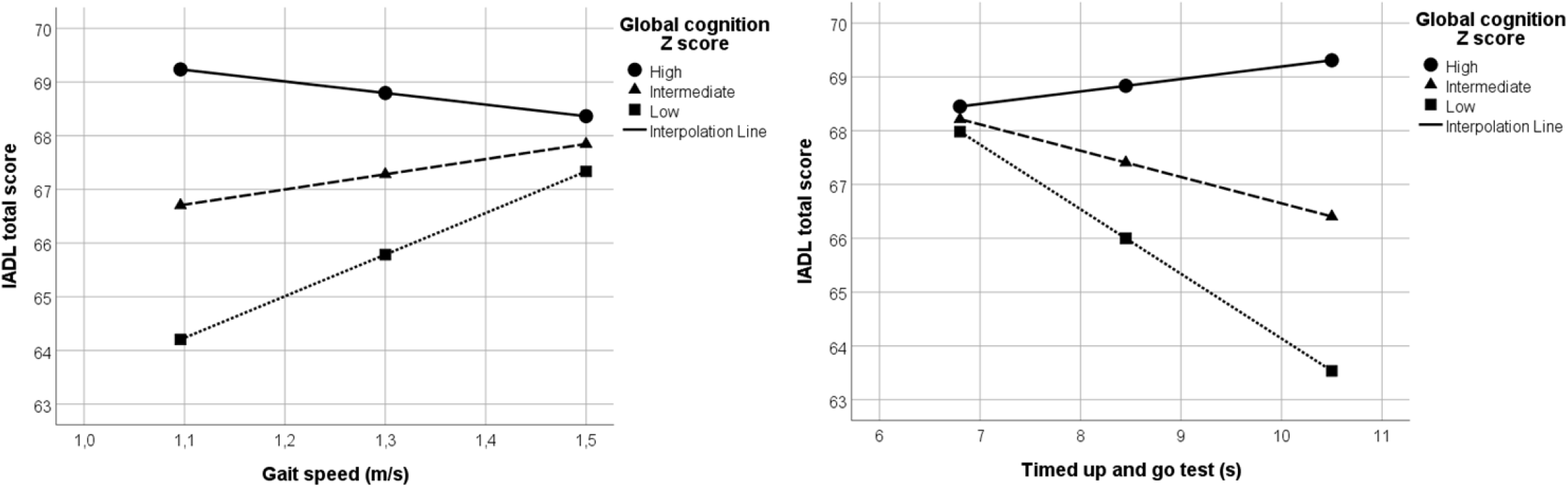
Combined associations of global cognitive and motor performances with instrumental activities on daily living (IADL) in subjects with WMH. Global cognition z score is depicted with interpolation lines according to 16^th^, 50^th^ and 84^th^ percentiles (low, intermediate and high performance, respectively) to show its potentiating effect on the associations between motor performances and IADL. Subjects with low performances in both cognitive and motor measures had the poorest outcome in IADL. The results of the interaction analyses are presented in table 5.

### Mediation analysis

Finally, we tested whether the cognitive and motor performances with the strongest relationships with IADL (global cognition and TUG) mediated the relationship of WMH or GM volume with IADL by using multivariable linear regression and the PROCESS macro for SPSS to estimate bootstrapped confidence intervals for the indirect effects.^29^ The direct effects between the variables are presented in tables 2 and 4 and the significant indirect effects in figure 2. Controlling for age, sex and education, the relationship between WMH and IADL was mediated by global cognition, which reduced the strength and thereby explained 28% of the association (standardized β for WMH decreased from -0.284 to -0.206). Global cognition also mediated the relationship between GM volume and IADL by 38% (standardized β for GM decreased from 0.239 to 0.149). Both indirect effects were significant: β -0.913, CI 95% -2.317–-0.065 and β 0.014, CI 95% 0.001–0.033, respectively (p<0.05). Performance in the TUG test significantly mediated the relationship between GM volume and IADL explaining 46% of the association (standardized β for GM decreased from 0.240 to 0.130; indirect effect β 0.016, CI 95% 0.004–0.033). However, the indirect effect of TUG on the association between WMH and IADL did not reach significance (>0.05).

**Figure 2.**
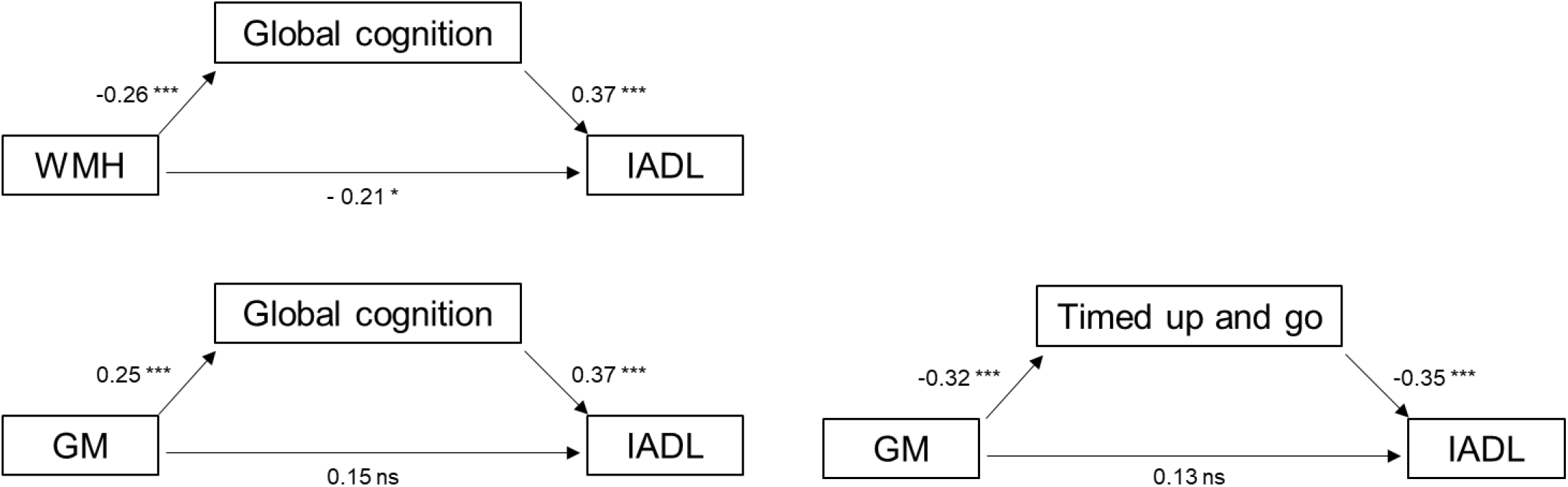
The relationship between white matter hyperintensities (WMH) and instrumental activities of daily living (IADL) was mediated by global cognition, whereas the relationship between gray matter (GM) volume and IADL was mediated by global cognition and timed up-and-go performance. Standardized β coefficients of linear regression analyses are presented adjacent to arrows (*** p<0.001; * p<0.05; ns, non-significant). The indirect effects were significant adjusting for age, sex and education.

## DISCUSSION

This study elaborated the complex relationship and clinical importance of cognitive and motor disturbances in older individuals with different degrees of SVD-related brain changes. We first confirmed the significance of WMH and GM volumes as determinants of motor, cognitive and functional impairments. We then showed that various aspects of motor and cognitive functions were strongly related to each other, and that combinations of motor and cognitive deficits had synergistic effects on functional impairment in IADL. Moreover, we demonstrated that the associations of WMH and GM with IADL were largely explained by related cognitive and motor deficits.

The association of WMH with reduced mobility in the elderly has been well established. The severity of WMH has been related to falls^11, 31^ as well as disturbances in gait, balance and functional mobility.^10, 13, 32^ In addition, brain atrophy as indicated by GM volume or cortical thinning has correlated with gait disturbances in SVD.^14, 15^ In this study, both WMH and GM volumes were significantly associated with various manifestations of motor disturbances such as impaired SPPB score, gait speed, balance and TUG performance, but not with self-reported recent falls. The associations of WMH and brain atrophy with cognitive decline in SVD has been also established in numerous studies.^5, 7, 33, 34^ Therefore expectedly, in the present sample, both of these interrelated brain changes were strongly linked with impairment in global cognition and in specific domains of executive functions, processing speed and memory.

Majority of previous studies have examined the cognitive and motor symptoms of SVD separately, so their mutual relationships have not been elucidated. To address this gap, we showed a consistent pattern of associations between various aspects of motor and cognitive functions independently of sociodemographic factors. Gait speed, balance and the TUG test reflecting functional mobility were most strongly associated with global cognition, executive functions and processing speed, whereas memory had weaker relations with motor abilities. Meta-analyses of studies with healthy elderly have suggested a close link between cognitive performance and risk of falls as well as gait dysfunction.^1, 35^ Of cognitive domains, executive functions have been most commonly connected with mobility.^1, 36, 37^ Very few studies have focused on the interrelations of cognition and mobility in patients with cerebrovascular disease. Recently, associations between measures of motor and cognitive abilities have been reported in older individuals with atrial fibrillation, although these relationships were not related to cerebral lesion burden.^17^ In chronic cerebrovascular disease, executive functions, but not global cognition, has been associated with risk of falls, while balance had no correlations with neuropsychological measures.^16^

The role of WMH in old-age disability in terms of impaired IADL has been well documented.^3^ We observed both WMH and GM volumes to be associated with IADL as evaluated with a validated questionnaire by the patient’s close informant. There were no significant relationships between the volumes and self-rated quality of life reflecting overall satisfaction to personal health, daily activities and life conditions. However, several measures of cognitive and motor functions were directly related to both IADL and quality of life, the strongest predictors being TUG and global cognition.

Of particular importance is that motor and cognitive deficits showed consistent synergistic interactions on IADL. Subjects with low scores in both cognitive and motor measures had remarkably poor IADL as compared to those with decline in only one function. This over and above effect was apparent for all cognitive (global, executive, speed, memory) and motor (gait, TUG, balance) measures, although the interactions with balance were somewhat weaker. To our knowledge, such findings have not been reported before in SVD. In a community sample, Rosano et al. have demonstrated an additive combined association of a processing speed test and gait speed with incident disability in basic activities of daily living, but the interaction between these measures did not reach significance.^38^

Furthermore, our results indicated that the association between WMH volume and IADL was mediated and thereby partly explained by global cognitive dysfunction. The association between GM volume with IADL, in turn, was mediated by functional mobility (TUG) and global cognition. Although age-related WMH and brain atrophy as well as cognitive and motor deficits are known to co-occur, these indirect pathways have not been statistically proven before. Interestingly however, the association between WMH volume and slower walking speed under dual-task conditions has been mediated in part by global cognition and executive abilities.^39^

Taken together, the results support the view that deterioration of motor and cognitive functions are interrelated processes and should not be seen as separate coincidental problems.^40^ Gait and cognitive deficits coexist already in the early stages of aging, are exacerbated by vascular pathomechanisms, and are important determinants of old-age disability.^40^ The interplay and synergism between cognitive and motor disturbances may reflect compromised integrity and disconnection in shared networks and brain regions susceptible to microvascular damage and vascular risk factors.

Our study is limited by the cross-sectional design, which precludes interpretations of causal relationships between brain changes, motor and cognitive abilities as well as functional outcome. Instead of analyzing the full variety of SVD-related brain changes,^9^ we focused on WMH and GM volumes shown to have the strongest associations with clinical outcomes.^5^ Since subjects with cortical infarcts and large subcortical infarcts were excluded from the study, there were only a few cases with small chronic infarctions evident on MRI. The age range of this study (65-75 years) provides a time window in which SVD brain changes and preclinical cognitive decline are already present, but the likelihood of coexisting neurodegenerative disorders is low. However, the results may not fully represent those of older age groups or more severe stages of vascular cognitive impairment. Moreover, 13 % of the subjects did not have an evaluation of IADL given by an informant. These subjects were comparable to the rests of the sample in age, education and WMH and GM volumes, but women were overrepresented, introducing possible selection bias.

The strengths of the study are the detailed evaluations of motor, cognitive and functional abilities with objective and well-validated tests. Neuropsychological assessment included multiple psychometrically sound tests within each domain to achieve robust composite measures for the most relevant cognitive domains. As has been recommended based on recent meta-analysis, a comprehensive test battery covering a range of cognitive domains is necessary to accurately assess the full extent of SVD-related cognitive impairments.^9^ In addition to age and sex, all analyses were controlled for years of education, which is an important indicator of individual cognitive reserve. However, we did not statistically control for vascular risk factors, since these were regarded as causal factors behind the pathogenesis of SVD and related brain changes.

In conclusion, this study highlights the combined effects of cognitive and motor impairments on functional outcome in covert SVD. Deficits in gait, balance and functional mobility had strong connections with global cognition, executive functions, processing speed and memory, and these dysfunctions together had synergistic detrimental associations with IADL. The findings also suggest that more severe WMH load and reduced GM volume contribute to disability in IADL through impaired global cognition and functional mobility. The results have profound clinical implications underlining the need for comprehensive approach in the assessment and management of SVD. Patients with both cognitive and motor impairments are at disproportionate risk for poor outcome and may benefit from multidomain interventions to promote functional independence.

## Supporting information

STROBE Statement

## Data Availability

Anonymized data will be shared on request from a qualified investigator.

---

**Supplementary Table 1.** Neuropsychological test battery of the Helsinki Small Vessel Disease Study.

| **Cognitive domain** | **Neuropsychological test** | **Variable** | **Reference** |
| --- | --- | --- | --- |
| Processing speed | WAIS-IV Coding | Correct responses in 120 sec | 1 |
|  | Stroop test (colour-congruent) | Correct responses in 45 sec | 2 |
|  | FAT Numbers (Trail making A) | Time | 3, 4 |
| Executive functions | Stroop test (colour-incongruent) | Correct responses in 45 sec | 2 |
|  | Verbal fluency test (phonemic and semantic) | Total number of correct responses | 5 |
|  | Hayling Sentence Completion Test | Error score | 6 |
|  | Brixton Spatial Anticipation Test | Error score | 6 |
|  | FAT Number-Letter (Trail making B) | Time / correct responses | 3, 4 |
| Working memory | WMS-III Letter-Number Sequencing | Correct score | 7 |
|  | WMS-III Digit Span | Forward and backward score | 7 |
|  | FAT Visuospatial Span | Forward and backward score | 3, 8 |
| Memory and learning | WMS-III Word Lists | Total immediate recall and delayed recall score | 7 |
|  | WMS-III Logical Memory | Immediate and delayed recall score | 7 |
|  | Rey-Osterrieth complex figure test | Delayed recall score | 5 |
| Visuospatial perception | WAIS-IV Block Design | Correct score | 1 |
|  | Rey-Osterrieth complex figure test | Copying score | 5 |
| Verbal reasoning | WAIS-IV Similarities | Correct score | 1 |

FAT; Flexible Attention Test (a computerised modification of the Trail making test and the Corsi block-tapping test), WAIS-IV, Wechsler Adult Intelligence Scale; WMS-III, Wechsler Memory Scale

